# Model-based reasoning methods for diagnosis in integrative medicine based on electronic medical records and natural language processing

**DOI:** 10.1101/2020.07.12.20151746

**Authors:** Wenye Geng, Xuanfeng Qin, Zhuo Wang, Qing Kong, Zihui Tang, Lin Jiang

## Abstract

**Background:** This study aimed to investigate model-based reasoning (MBR) algorithms for the diagnosis of integrative medicine based on electronic medical records (EMRs) and natural language processing.

**Methods:** A total of 14,075 medical records of clinical cases were extracted from the EMRs as the development dataset, and an external test dataset consisting of 1,000 medical records of clinical cases was extracted from independent EMRs. MBR methods based on word embedding, machine learning, and deep learning algorithms were developed for the automatic diagnosis of syndrome pattern in integrative medicine. MBR algorithms combining rule-based reasoning (RBR) were also developed. A standard evaluation metrics consisting of accuracy, precision, recall, and F1 score were used for the performance estimation of the methods. The association analyses were conducted on the sample size, number of syndrome pattern type, and diagnosis of lung diseases with the best algorithms.

**Results:** The Word2Vec CNN MBR algorithms showed high performance (accuracy of 0.9586 in the test dataset) in the syndrome pattern diagnosis. The Word2Vec CNN MBR combined with RBR also showed high performance (accuracy of 0.9229 in the test dataset). The diagnosis of lung diseases could enhance the performance of the Word2Vec CNN MBR algorithms. Each group sample size and syndrome pattern type affected the performance of these algorithms.

**Conclusion:** The MBR methods based on Word2Vec and CNN showed high performance in the syndrome pattern diagnosis in integrative medicine in lung diseases. The parameters of each group sample size, syndrome pattern type, and diagnosis of lung diseases were associated with the performance of the methods.

**Strengths and limitations of this study:**

1. A novel application of artificial intelligence – natural language processing approaches on diagnosis of integrative medicine
2. A study of medical artificial intelligence based on real-world data of electronic medical records
3. Multiple approaches on artificial intelligence to include traditional machine learning algorithms, neural network, and deep learning algorithms
4. Rule-based combining model-based reasoning to be explored in this dataset

## Background

Integrative medicine is a medical form that combines practices and treatments from alternative medicine with conventional medicine [1-3]. In China, integrative medicine combines traditional Chinese medicine (TCM) and modern medicine for clinical practice [1-3]. The diagnosis of integrative medicine comprises the clinical diagnosis of modern medicine and syndrome pattern diagnosis [4]. Syndrome pattern based on TCM theory is an outcome of the analysis of TCM information by the TCM practitioner, and TCM treatments rely on it [4]. A syndrome pattern can be defined as a categorized pattern of symptoms and signs in a patient at a specific stage during the course of a disease. Syndrome elements are the smaller units of syndrome classification and the basic elements of a syndrome pattern [5]. The correct combination of syndrome elements can infer an appropriate syndrome pattern. Syndrome elements are also derived from the syndrome and signs from the patient [5, 6]. Generally, practitioners of integrative medicine making diagnosis decisions need to combine syndrome pattern diagnosis and the diagnosis of modern medicine [5, 6]. As TCM treatments rely on syndrome pattern diagnosis, the treatment combined with the therapies of TCM and modern medicine is more efficient for patients. Therefore, syndrome pattern for the diagnosis of integrative medicine is an essential part of diagnosis.

Electronic medical records (EMRs) are the systematized collection of patients’ and the population’s electronically stored health information in a digital format that can be shared across different healthcare settings [7, 8]. In China, EMRs are a collection of diagnoses of syndrome patterns and model medicine as well as syndromes and signs with the TCM format [7, 8]. Natural language processing (NLP) is a field of artificial intelligence and computational linguistics concerned with the interactions between computers and human natural languages [9, 10]. Currently, NLP techniques combining EMRs have been comprehensively applied to medical data mining and medical decision support system [9, 10]. Word embedding, as one of the techniques in NLP, attempted to map a word using a dictionary to a vector of real numbers in a low-dimensional space [11, 12]. It is important in EMR data mining or artificial intelligence application in medicine for medical texts to be transferred to vectors because computers can handle or understand medical texts through computability vectors.

Applying artificial intelligence techniques to support physicians in medical practices is a major challenge. The processing of uncertainty information mainly contributes to the challenge. Syndrome and sign information is under the classic uncertainty information. The artificial neural network (ANN) can successfully and efficiently handle syndrome and sign information with uncertainty [13]. ANN is a computational model based on the structure and functions of biological neural networks [14]. The remarkable information processing characteristics of the ANN in terms of nonlinearity, fault and noise tolerance, high parallelism, and learning and generalization capabilities contribute to uncertain information processing and quantitative analysis. Furthermore, model-based reasoning (MBR) methods based on machine learning or ANN can successfully process syndrome and sign information with uncertainty to make a precise and accurate diagnosis of integrative medicine.

As mentioned previously, syndrome and sign information or relative information can be extracted from the EMRs, and content texts can be mapped to computability vectors using NLP techniques. Furthermore, MBR methods can be used to create a computer-aided system to support the diagnosis of integrative medicine. However, only a few studies have been conducted on MBR methods with EMRs and NLP to support the diagnosis of integrative medicine. Fortunately, our previous work was carried out to analyze syndrome patterns and syndrome elements in lung diseases based on real-world EMR data [5]. This study aimed to explore MBR algorithms in the diagnosis of integrative medicine based on EMRs and NLP techniques in lung disease datasets. We also estimated the associations among the factors of sample size, number of syndrome pattern type, and diagnosis of modern medicine using the MBR algorithms.

## Methods

The workflow of the analysis of the MBR methods in the diagnosis of integrative medicine based on EMRs and NLP is illustrated in Figure 1. The EMRs on lung diseases were exported from the hospital information system, and the syndrome and sign information and relative information were extracted as a text format. The corresponding syndrome pattern diagnosis, clinical diagnosis of modern medicine, and syndrome elements were extracted and saved to the database with the structure data according to the unique code of patients. The content texts of the syndrome and sign information were mapped to the computability vectors through word embedding. The classification models that include the vectors of syndrome and sign information and syndrome patterns or syndrome elements were developed using machine learning or neural network methods. MBR algorithms were developed on the basis of classification models concerning the syndrome pattern, and the model-based and rule reasoning algorithms were developed using the classification models and rule knowledge based on the combination of syndrome elements and syndrome patterns. The performances of the MBR methods in the diagnosis of integrative medicine in lung diseases evaluated and compared (for the main program codes for the module, please see https://github.com/zihuitang/clincial_decision_support_system_im).

**Figure 1:**
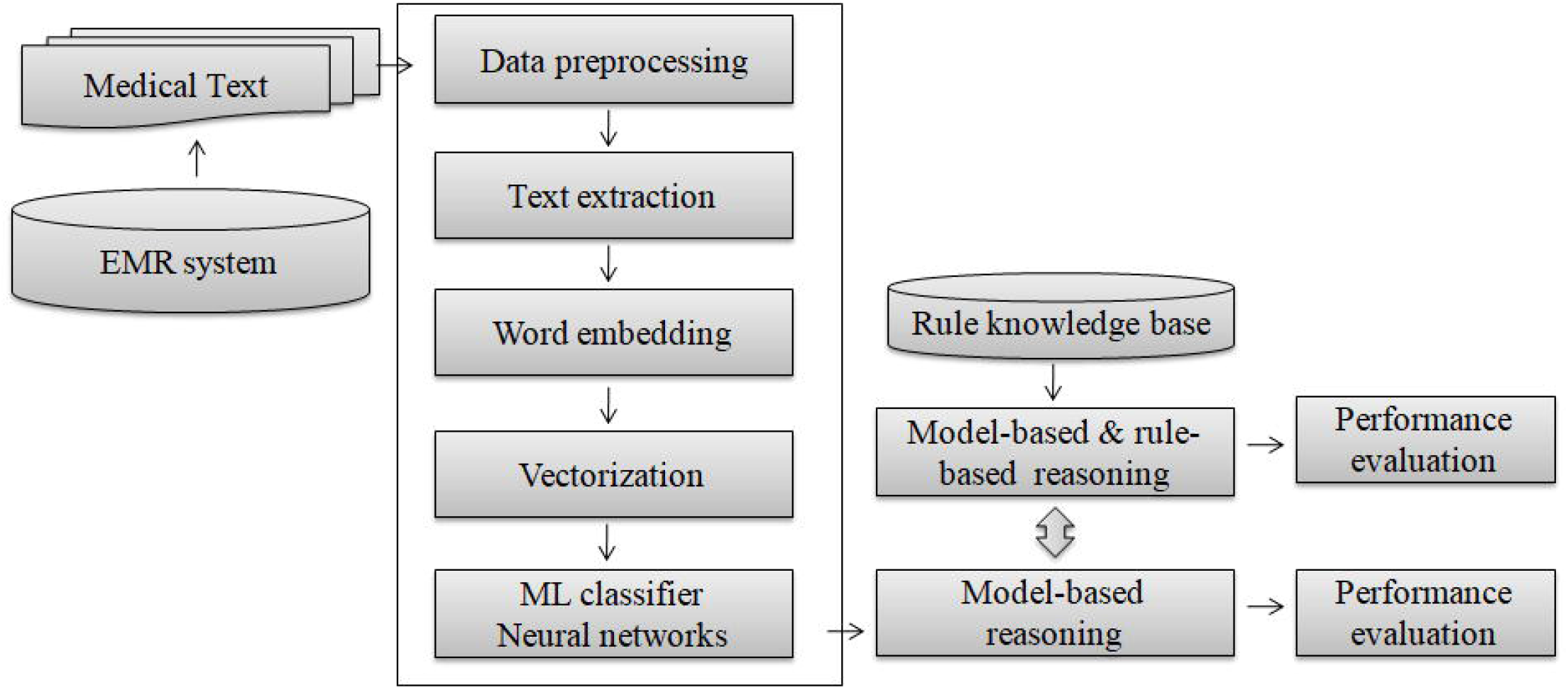
The workflow of model based reasoning methods development on diagnosis in integrative medicine based on electronic medical records with natural language processing.

### Data collection and processing

In our previous real-world study on the syndrome pattern and syndrome element of lung disease, EMRs were collected from lung disease wards in five hospitals [5]. A dataset consisting of 14,075 medical records of clinical cases from four hospitals was assigned as the development dataset, and it was divided into the train dataset and the test dataset at a ratio of 4:1. Another independent dataset comprising 1,000 medical records of clinical cases from a hospital was set as the external test dataset. The information comprised patients’ identity number, ward number, admission time, admission notes, first medical records, general medical records, discharge note, diagnosis of syndrome pattern, and diagnosis of modern medicine. In this work, we selected 10 common syndrome pattern types and 8 common lung diseases in the lung disease wards. Nine syndrome element types were generated and combined with the corresponding 10 syndrome pattern types.

### Medical information extraction

The Chinese text information on the chief complaints, syndromes, and positive signs in the chest, tongue, and pulse was extracted from the admission notes, first medical records, and discharge records (Figure 2). The extracted Chinese text information was combined into contexts called four diagnoses in TCM. The contexts of the syndromes and signs underwent word-cutting process to split them into tokens. In this work, the first corpus included the context of syndrome and sign information. In the analysis of the association diagnosis of modern medicine and syndrome pattern diagnosis, another corpus included an additional token of diagnosis of modern medicine.

**Figure 2:**
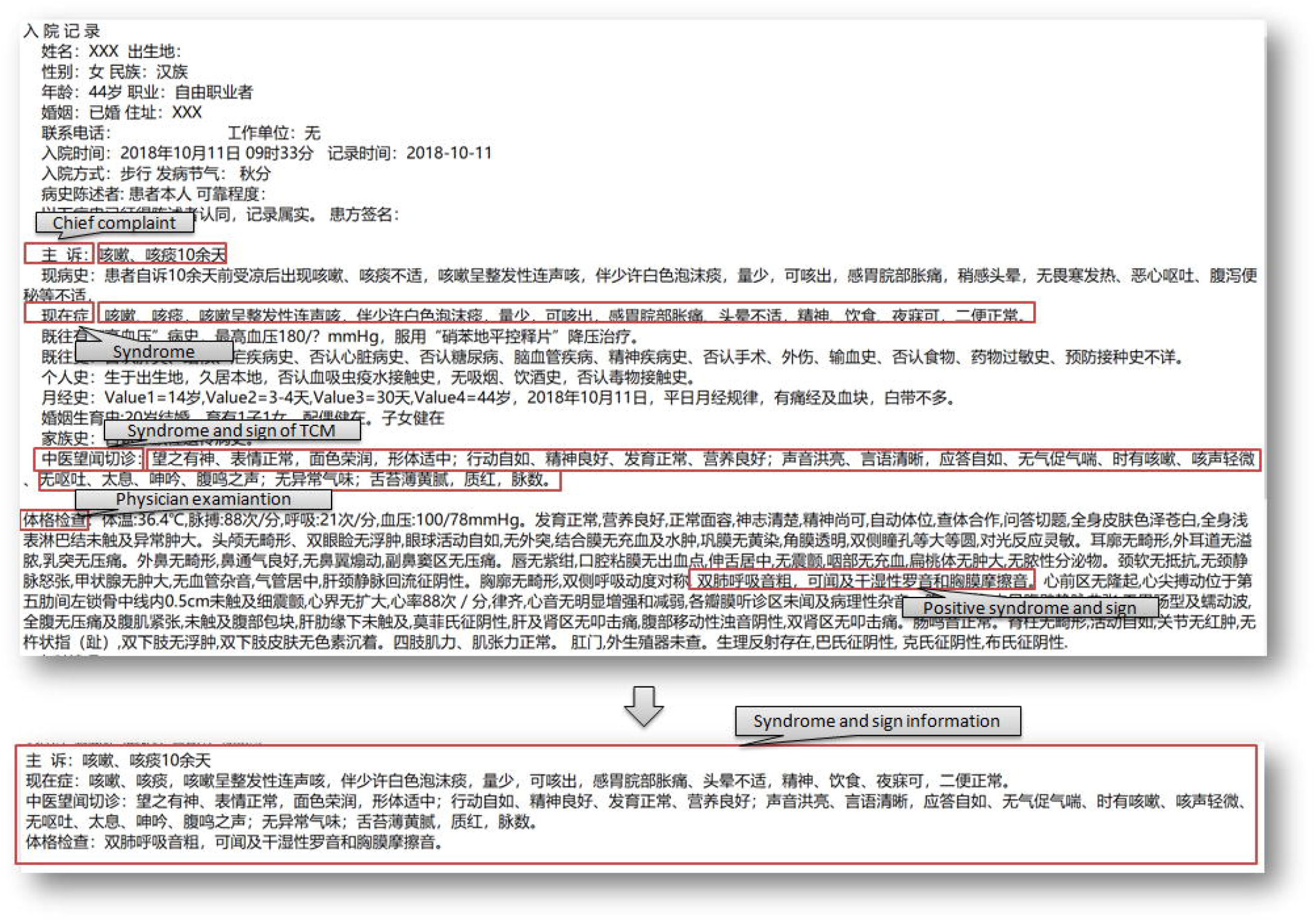
Syndrome and sign information extracted from electronic medical records on integrative medicine in lung disease.

### Word2Vec

Word embedding is an NLP feature-learning technique in which words are mapped to vectors of real numbers [15]. Word embedding involves mathematical embedding from a space with one dimension per word to a continuous vector space with a much lower number of dimensions. The Word2Vec model is an NLP system that is used to produce word embedding, which takes a large corpus of text as its input and produces a vector space, and each unique word in the corpus is assigned a corresponding vector in the space [15]. The Word2Vec model generates vectors for each word present in a document. In this study, the corpus from a Chinese language Wikipedia dump, which is available at https://dumps.wikimedia.org/zhwiki/latest/zhwiki-latest-pages-articles.xml.bz2, was used to pre-train the word vector model. The parameters utilized with the word2Vec model were developed for dimension reduction into 256 dimension vectors, 5 context windows, and a minimum sentence word count of 10. The Word2Vec model was implemented using the Gensim Python library [16].

### Doc2Vec

The Doc2Vec model is an extension of Word2Vec that constructs embeddings from entire documents or sentences (instead of individual words) to learn a randomly initialized vector for the document (or sentence) along with the words [17]. The Doc2Vec model modifies the Word2Vec algorithm into an unsupervised learning algorithm that produces continuous representations for large blocks of texts, such as sentences, paragraphs, or entire documents. In this work, Doc2Vec was used to produce vectors for texts. The corpus from a Chinese language Wikipedia dump was again used to pre-train the doc vector model. The parameters utilized with the Doc2Vec model were developed in the dimension reduction into 192 dimension vectors, 5 context windows, and a minimum sentence word count of 10. The Doc2Vec model was also implemented using the Gensim Python library.

### Machine learning

In this work, the four different machine learning classifiers algorithms, namely, random forest (RF), extreme gradient boosting (XGBoost), support vector machines (SVM), and K-nearest neighbor (KNN), were used to develop MBR [18-20]. RF, a classic machine learning classifier, is composed of tree predictors, with each tree depending on the values of a random vector sampled independently and having the same distribution for all trees in the forest [21]. RF aims to reduce the tree correlation issue by choosing only a subsample of the feature space at each split. In this work, RF was used on 1,000 trees in the forest, and it was implemented using the scikit-learn Python library.

XGBoost is an optimized distributed gradient-boosting system designed to be highly efficient, flexible, and portable [22]. It implements machine learning algorithms under the gradient boosting framework, which attempts to accurately predict a target variable by combining an ensemble of estimates from a set of simpler, weaker models. XGBoost can also be implemented using the scikit-learn Python library.

SVM is a well-known supervised learning model associated with learning algorithms that analyze data used for classification and regression analysis [23]. SVM was useful in text-based classification tasks and to not be prone to error in high-dimensional datasets. In this work, SVM was used with a linear kernel and implemented using the scikit-learn Python library.

The KNN classifier, one of the most popular machine learning algorithms, is based on the Euclidean distance between a test sample and the specified training samples [24]. It is used for data classification that attempts to determine in which group a data point is included by examining the data points around it. In this study, KNN was implemented using the scikit-learn Python library.

### Artificial neural network

ANNs, one of the main tools used in machine learning, are a group of models inspired by biological neural networks used for estimating functions that depend on a large number of inputs [13]. ANN algorithms have two different classifiers: multilayer perceptron (MLP) and convolutional neural network (CNN). MLP is a feed-forward ANN model that maps sets of input data onto a set of appropriate outputs [25]. It consists of multiple layers of nodes with a nonlinear activation function in a directed graph, with each layer fully connected to the next one. Back-propagation is used as a supervised learning technique in MLP. In this work, MLP was performed with six hidden layers, with the nodes per layer varying from 64 to 1024. It was also implemented using the scikit-learn Python library.

CNN is one of the most popular algorithms for deep learning [26]. It is a category of ANN in which a model learns to perform classification tasks directly from images, text, or sound, and it has been proven effective in the areas of text classification and image recognition. CNN comprises one or more convolutional layers with a sub-sampling step, followed by one or more fully connected layers as in a standard multilayer neural network [27]. In this work, CNN was performed with an embedding layer, a convolutional layer, a max pooling layer, and two fully connected layers, and it was implemented using the Keras Python library.

### MBR

In this study, the development of MBR was based on word embedding and machine learning classifiers for syndrome pattern [28, 29]. A total of 11 MBR algorithms were used: Word2Vec RF, Word2Vec XGBoost, Word2Vec SVM, Word2Vec KNN, Word2Vec MLP, Word2Vec CNN, Doc2Vec RF, Doc2Vec XGBoost, Doc2Vec SVM, Doc2Vec KNN, and Doc2Vec MLP. These models with multiclass outputs were consistent with the syndrome pattern types. A comparison of the performance of the 11 MBR algorithms was conducted.

### MBR combined with rule-based reasoning (RBR)

MBR was based on word embedding and machine learning classifiers for syndrome elements. Nine MBR algorithms were used: Word2Vec RF, Word2Vec XGBoost, Word2Vec KNN, Word2Vec MLP, Word2Vec CNN, Doc2Vec RF, Doc2Vec XGBoost, Doc2Vec KNN, and Doc2Vec MLP. These models with multi-label outputs were consistent with the syndrome element types. The syndrome patterns were generated by combining the syndrome elements, which follow the rule knowledge base of the syndrome elements, with the syndrome pattern. A comparison of the performance of the nine MBR combined with RBR algorithms was performed.

### Evaluation

The performances of the MBR algorithms in syndrome pattern were evaluated in test dataset and the external dataset using standard metrics consisting of accuracy, precision, recall, and F1-score [30]. Moreover, the performances of the Word2Vec CNN MBR algorithms in each syndrome pattern and each syndrome element were evaluated in test dataset by using standard metrics. A five-fold cross-validation was conducted 20 times on the train dataset for each algorithm to estimate the 95% confidence interval (CI) for the performance parameters.

The accuracy comparison analysis of the Word2Vec CNN MBR algorithms in corpus 1 and corpus 2 were conducted in different proportions of the sample size of the development dataset. In the accuracy analysis of the dataset, each group sample size was set as proportion of total sample size and the number of syndrome pattern type were selected randomly. The linear regression analyses were conducted to evaluate the associations between each group sample size and the number of syndrome pattern type at accuracies of 0.90 and 0.95 of the methods.

## Results

### Development and external datasets

The characteristics of the dataset are shown in Figure 3. The development dataset consisted of 14,075 medical records of clinical cases, and the external dataset had 1,000 medical records of clinical cases. Eight common lung diseases were found in the development dataset: lung cancer (18.42%), pulmonary infection (18.59%), acute bronchitis (8.39%), interstitial pneumonia (1.66%), chronic bronchitis (9.78%), chronic obstructive pulmonary disease (COPD, 25.98%), bronchiectasis (4.31%), and asthma (12.88%) (Figure 3A). The same common lung diseases with the same proportions were also found in the external dataset (Figure 3B). Ten common syndrome pattern types were found in the development dataset: qi-deficiency of lung and spleen, qi-deficiency of lung and kidney, yin-deficiency of lung, wind-cold attacking lung, wind-heat attacking lung, cold wheezing, deficiency of qi and yin, hot wheezing, phlegm-heat obstruction in lung, and phlegm obstruction in lung (Figure 3C). The same 10 syndrome pattern types with the same proportions were found in the external dataset (Figure 3D). The development dataset had 35,992 syndrome elements for 14,075 syndrome patterns, and a syndrome pattern consisted of 2.56 syndrome elements on average. The development dataset included nine syndrome element types: phlegm, wind, cold, heat, qi-deficiency, yin-deficiency, lung, spleen, and kidney (Figure 3E).A total of 2,602 syndrome elements with the same nine types were found in 1,000 syndrome patterns (Figure 3F).

**Figure 3:**
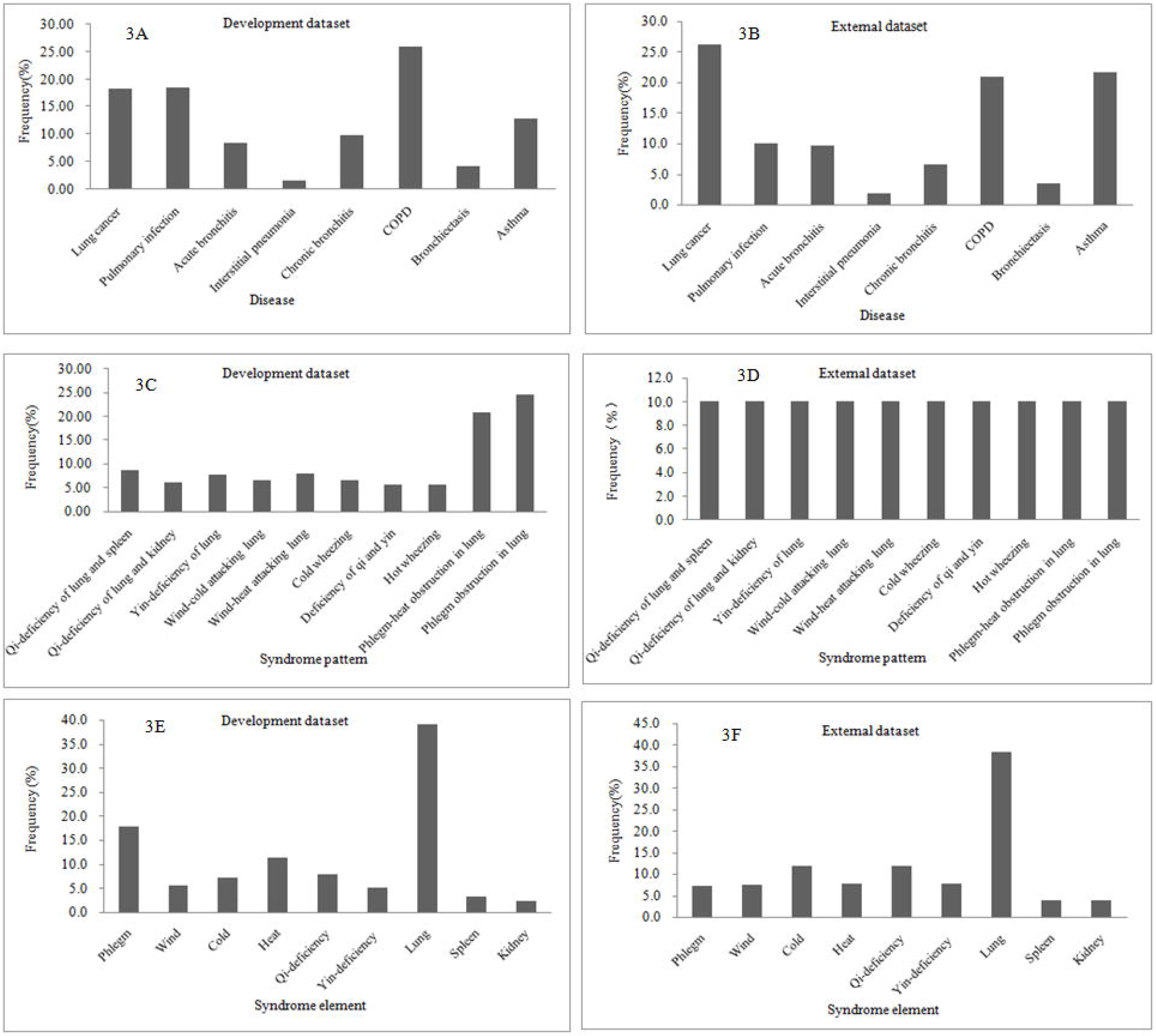
Distribution of clinical diagnosis, syndrome pattern, syndrome element on lung disease in development dataset and external test dataset.

### MBR

In the test dataset, the performance analysis of the MBR based on Word2Vec to identify syndrome patterns showed an average accuracy of 0.9397 (95%CI: 0.9312–0.9468) in the Word2Vec RF model and 0.9323 (95%CI: 0.9213–0.9443) in the Word2Vec ANN model (Table 1). The highest average accuracy was 0.9471 (95%CI: 0.9382–0.9549) in the Word2Vec CNN model. The parameters of precision, recall, and F1 score were 0.9478 (95%CI: 0.9393–0.9557), 0.9471 (95%CI: 0.9382–0.9549), and 0.9470 (95%CI: 0.9383 – 0.9550) in the Word2Vec CNN model, respectively. Similar performance values were found in the corresponding external dataset.

**Table 1:**
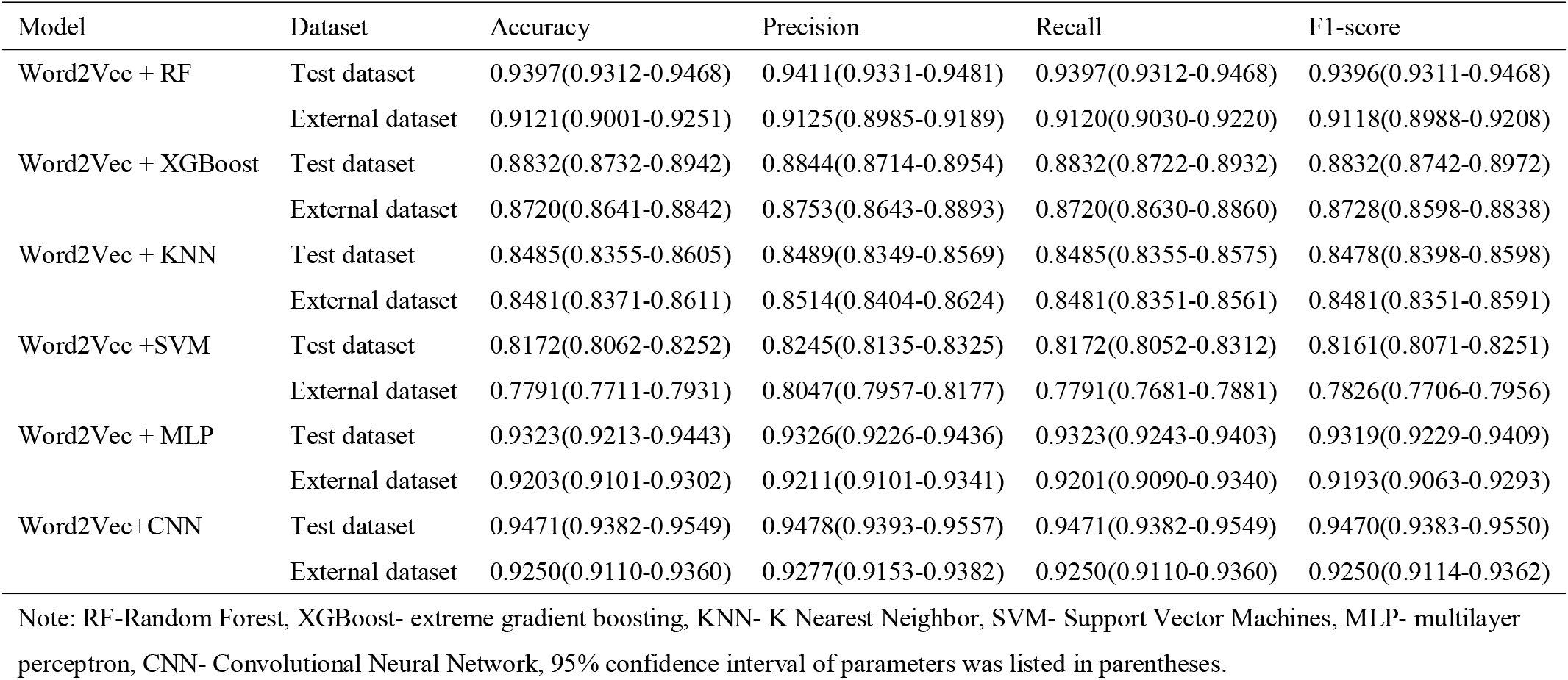
Performance analysis of model-based reasoning methods on syndrome pattern diagnosis for lung disease based on Word2Vec in test dataset and external dataset

The performance analysis of the MBR based on Doc2Vec to identify syndrome patterns in the test dataset showed the highest average accuracy of0.8840 (95%CI: 0.8730–0.8970) in the Word2Vec CNN model (Table 2). The parameters of precision, recall, and F1 score were 0.8876 (95%CI: 0.8776–0.8976), 0.8840 (95%CI: 0.8710–0.8932), and 0.8843 (95%CI: 0.8753–0.8973) in the Doc2Vec CNN model, respectively. Similar performance values were found in the corresponding external dataset.

**Table 2:** Performance analysis of model based reasoning methods on syndrome pattern diagnosis for lung disease based on Doc2Vec in test dataset and external dataset

| Model | Dataset | Accuracy | Precision | Recall | F1-score |
| --- | --- | --- | --- | --- | --- |
| Doc2Vec + RF | Test dataset | 0.8320(0.8198-0.8442) | 0.8457(0.8345-0.8567) | 0.8320(0.8198-0.8442) | 0.8337(0.8217-0.8458) |
|  | External dataset | 0.8190(0.8090-0.8310) | 0.8506(0.8366-0.8610) | 0.8190(0.8110-0.8323) | 0.8267(0.8147-0.8397) |
| Doc 2Vec + XGBoost | Test dataset | 0.7584(0.7444-0.7724) | 0.7682(0.7602-0.7812) | 0.7584(0.7504-0.7704) | 0.7589(0.7499-0.7719) |
|  | External dataset | 0.7270(0.719-0.7400) | 0.7735(0.7645-0.7835) | 0.7270(0.7130-0.7390) | 0.7391(0.7261-0.7501) |
| Doc 2Vec + KNN | Test dataset | 0.8527(0.8407-0.8637) | 0.8588(0.8488-0.8668) | 0.8527(0.8407-0.8627) | 0.8535(0.8425-0.8665) |
|  | External dataset | 0.8202(0.8092-0.8282) | 0.8246(0.8116-0.8326) | 0.8220(0.8090-0.8331) | 0.8215(0.8105-0.8295) |
| Doc 2Vec +SVM | Test dataset | 0.6748(0.6628-0.6848) | 0.7424(0.7334-0.7504) | 0.6748(0.6668-0.6858) | 0.7577(0.7467-0.7667) |
|  | External dataset | 0.5820(0.5700-0.5950) | 0.5743(0.5663-0.5883) | 0.5920(0.5830-0.6033) | 0.5288(0.5168-0.5388) |
| Doc 2Vec + MLP | Test dataset | 0.8840(0.8730-0.8970) | 0.8876(0.8776-0.8976) | 0.8840(0.8710-0.8932) | 0.8843(0.8753-0.8973) |
|  | External dataset | 0.8760(0.8620-0.8890) | 0.8897(0.8757-0.9027) | 0.8760(0.8630-0.8851) | 0.8791(0.8701-0.8921) |

Note: RF-Random Forest, XGBoost- extreme gradient boosting, KNN- K Nearest Neighbor, SVM- Support Vector Machines, MLP- multilayer perceptron, 95% confidence interval of parameters was listed in parentheses.

### MBR combined with RBR

The performance analysis of the MBR combined with RBR based on Word2Vec in the test dataset reported that the highest average accuracy was 0.9229 (95%CI: 0.9099–0.9319) in the word2Vec CNN model (Table 3). The parameters of precision, recall, and F1 score were 0.9884 (95%CI: 0.9744–0.9964), 0.9679 (95%CI: 0.9589–0.9809), and 0.9778 (95%CI: 0.9698–0.9888) in the Word2Vec CNN model, respectively. Similar performance values were found in the corresponding external dataset.

**Table 3:** Performance analysis of models based reasoning combining rule based reasoning methods on syndrome pattern diagnosis for lung disease based on Word2Vec in test dataset and external dataset

| Model | Dataset | Accuracy | Precision | Recall | F1-score |
| --- | --- | --- | --- | --- | --- |
| Word2Vec + RF | Test dataset | 0.9131(0.8990-0.9261) | 0.9934(0.9814-0.9983) | 0.9628(0.9538-0.9748) | 0.9774(0.9644-0.9864) |
|  | External dataset | 0.9040(0.8903-0.9180) | 0.9657(0.9547-0.9747) | 0.9580(0.9501-0.9721) | 0.9617(0.9477-0.9697) |
| Word2Vec + XGBoost | Test dataset | 0.7703(0.7583-0.7803) | 0.9666(0.9556-0.9786) | 0.9044(0.8924-0.9144) | 0.9333(0.9233-0.9433) |
|  | External dataset | 0.7980(0.7871-0.8112) | 0.9702(0.9582-0.9812) | 0.9227(0.9137-0.9337) | 0.9444(0.9364-0.9544) |
| Word2Vec + KNN | Test dataset | 0.8414(0.8324-0.8534) | 0.9380(0.9270-0.9502) | 0.9254(0.9164-0.9334) | 0.9312(0.9202-0.9432) |
|  | External dataset | 0.8521(0.8403-0.8612) | 0.9441(0.9321-0.9571) | 0.9373(0.9263-0.9473) | 0.9446(0.9306-0.9556) |
| Word2Vec + MLP | Test dataset | 0.9052(0.8930-0.9181) | 0.9751(0.9621-0.9830) | 0.9758(0.9678-0.9858) | 0.9752(0.9652-0.9862) |
|  | External dataset | 0.9021(0.8940-0.9151) | 0.9791(0.9671-0.9911) | 0.9780(0.9660-0.9904) | 0.9784(0.9704-0.9904) |
| Word2Vec+CNN | Test dataset | 0.9229(0.9099-0.9319) | 0.9884(0.9744-0.9964) | 0.9679(0.9589-0.9809) | 0.9778(0.9698-0.9888) |
|  | External dataset | 0.9160(0.9030-0.9261) | 0.9765(0.9655-0.9885) | 0.9662(0.9582-0.9782) | 0.9698(0.9608-0.9778) |

Note: RF-Random Forest, XGBoost- extreme gradient boosting, KNN- K Nearest Neighbor, MLP- multilayer perceptron, CNN- Convolutional Neural Network, 95% confidence interval of parameters was listed in parentheses.

The performance analysis of the MBR combined with RBR based on Doc2Vec showed that the highest average accuracy was 0.8190 (95%CI: 0.8082–0.8281) in the Doc2Vec CNN model (Table 4). The parameters of precision, recall, and F1 score were0.9550 (95%CI: 0.9441–0.9673), 0.9507 (95%CI: 0.9387–0.9597), and 0.9524 (95%CI: 0.9444–0.9654) in the Doc2Vec CNN model, respectively. Similar performance values were found in the corresponding external dataset.

**Table 4:** Performance analysis of models based reasoning combining rule based reasoning methods on syndrome pattern diagnosis for lung disease based on Doc2Vec in test dataset and external dataset

| Model | Dataset | Accuracy | Precision | Recall | F1-score |
| --- | --- | --- | --- | --- | --- |
| Doc2Vec + RF | Test dataset | 0.6410(0.6281-0.6520) | 0.8586(0.8496-0.8698) | 0.9745(0.9635-0.9865) | 0.9049(0.8939-0.9139) |
|  | External dataset | 0.5940(0.5810-0.6061) | 0.9728(0.9648-0.9828) | 0.8002(0.7892-0.8112) | 0.8642(0.8542-0.8762) |
| Doc 2Vec + XGBoost | Test dataset | 0.6177(0.6087-0.6307) | 0.8525(0.8415-0.8625) | 0.9413(0.9273-0.9513) | 0.8891(0.8771-0.8981) |
|  | External dataset | 0.536(0.5272-0.5440) | 0.9346(0.9266-0.9486) | 0.7863(0.7763-0.7953) | 0.8401(0.8301-0.8531) |
| Doc 2Vec + KNN | Test dataset | 0.8488(0.8358-0.8618) | 0.9393(0.9283-0.9523) | 0.9503(0.9383-0.9613) | 0.9440(0.9331-0.9582) |
|  | External dataset | 0.8260(0.8174-0.8383) | 0.9203(0.9073-0.9323) | 0.9415(0.9275-0.9535) | 0.9301(0.9211-0.9401) |
| Doc 2Vec + MLP | Test dataset | 0.8190(0.8082-0.8281) | 0.9550(0.9441-0.9673) | 0.9507(0.9387-0.9597) | 0.9524(0.9444-0.9654) |
|  | External dataset | 0.8031(0.7911-0.8111) | 0.9478(0.9398-0.9618) | 0.9446(0.9316-0.9546) | 0.9444(0.9314-0.9544) |

Note: RF-Random Forest, XGBoost- extreme gradient boosting, KNN- K Nearest Neighbor, MLP- multilayer perceptron, CNN- Convolutional Neural Network, 95% confidence interval of parameters was listed in parentheses.

### Word2Vec CNN MBR in corpus 1 and corpus 2

Corpus 1 included the syndrome and sign information without a clinical diagnosis of lung disease, whereas corpus 2 included the syndrome and sign information with a clinical diagnosis of lung disease. A higher average accuracy (0.9584, 95% CI: 0.9510–0.9655) was found in the Word2Vec CNN model in syndrome pattern diagnosis in corpus 2 than in corpus 1 (0.9471, 95% CI: 0.9382–0.9549) in the test dataset (Table 5). Moreover, higher performance parameter values of precision, recall, and F1 score were found in the Word2Vec CNN model in each syndrome pattern diagnosis in corpus2 than in corpus 1 (Table 5). Similar results were found in the Word2Vec CNN combined with RBR model in syndrome pattern diagnosis in corpus 2 in comparison with the model in corpus 1 in the test dataset with a full sample size (Table 6). A higher average accuracy of the Word2Vec CNN model was found in syndrome pattern diagnosis in the test dataset with different sample sizes in corpus 2 than in corpus 1 (Figure 4).

**Table 5:** Performance analysis of model based reasoning methods for each syndrome pattern in test dataset with Corpus 1 and Corpus 2

| Syndrome pattern | Corpus 1 |  |  |  | Corpus 2 |  |  |  |
| --- | --- | --- | --- | --- | --- | --- | --- | --- |
|  | Precision | Recall | F1 score | Support | Precision | Recall | F1 score | Support |
| Qi-deficiency of lung and spleen | 0.9363 | 0.9514 | 0.9438 | 247 | 0.9957 | 0.9665 | 0.9809 | 239 |
| Qi-deficiency of lung and kidney | 0.9362 | 0.9999 | 0.9670 | 176 | 0.9781 | 0.9944 | 0.9861 | 179 |
| Yin-deficiency of lung | 0.9777 | 0.9733 | 0.9755 | 225 | 0.9902 | 0.9999 | 0.9951 | 203 |
| Wind-cold attacking lung | 0.9943 | 0.9943 | 0.9956 | 176 | 0.9878 | 0.9999 | 0.9939 | 162 |
| Wind-heat attacking lung | 0.9899 | 0.9120 | 0.9494 | 216 | 0.9150 | 0.9826 | 0.9476 | 230 |
| Cold wheezing | 0.9724 | 0.9832 | 0.9778 | 179 | 0.9750 | 0.9653 | 0.9701 | 202 |
| Deficiency of qi and yin | 0.9934 | 0.9804 | 0.9868 | 153 | 0.9932 | 0.9932 | 0.9945 | 147 |
| Hot wheezing | 0.9051 | 0.9931 | 0.947 | 144 | 0.9563 | 0.9808 | 0.9684 | 156 |
| Phlegm-heat obstruction in lung | 0.9389 | 0.9021 | 0.9201 | 613 | 0.9357 | 0.9125 | 0.9240 | 606 |
| Phlegm obstruction in lung | 0.9183 | 0.9344 | 0.9263 | 686 | 0.9461 | 0.9407 | 0.9434 | 691 |
| Average (weighted) | 0.9477 | 0.9471 | 0.9470 | 2815 | 0.9586 | 0.9584 | 0.9584 | 2815 |

Note: Corpus 1 was consisting of syndrome and sign information, Corpus 2 was consisting of syndrome and sign information plus clinical diagnosis information. The average accuracy was 0.9471 (95% CI: 0.9382-0.9549) for syndrome pattern in test dataset with Corpus 1, and 0.9584 (95% CI: 0.9510-0.9655) for syndrome pattern in test dataset with Corpus 2.

**Table 6:** Performance analysis of model based reasoning combing rule based reasoning methods for each syndrome element in test dataset with Corpus 1 and Corpus 2

| Syndrome element | Corpus 1 |  |  |  | Corpus 2 |  |  |  |
| --- | --- | --- | --- | --- | --- | --- | --- | --- |
|  | Precision | Recall | F1 score | Support | Precision | Recall | F1 score | Support |
| Phlegm | 0.9907 | 0.9538 | 0.9719 | 1233 | 0.9935 | 0.9951 | 0.9943 | 1233 |
| Wind | 0.9926 | 0.9218 | 0.9559 | 435 | 0.9953 | 0.9770 | 0.9861 | 435 |
| Cold | 0.9800 | 0.9722 | 0.976 | 503 | 0.996 | 1.000 | 0.998 | 503 |
| Heat | 0.9704 | 0.8903 | 0.9286 | 811 | 0.9675 | 0.9174 | 0.9418 | 811 |
| Qi-deficiency | 0.9616 | 0.9756 | 0.9686 | 616 | 0.9871 | 0.9935 | 0.9903 | 616 |
| Yin-deficiency | 1.000 | 0.9851 | 0.9925 | 403 | 0.9975 | 0.9801 | 0.9887 | 403 |
| Lung | 1.000 | 1.000 | 1.000 | 2815 | 1.000 | 1.000 | 1.000 | 2815 |
| Spleen | 0.9644 | 0.9457 | 0.955 | 258 | 0.9771 | 0.9922 | 0.9846 | 258 |
| Kidney | 0.9882 | 0.9825 | 0.9853 | 171 | 0.9826 | 0.9883 | 0.9854 | 171 |
| Average (weighted) | 0.9885 | 0.968 | 0.9779 | 7245 | 0.9922 | 0.9863 | 0.9892 | 7245 |

Note: Corpus 1 consisting of syndrome and sign information, Corpus 2 consisting of syndrome and sign information plus clinical diagnosis information. The average accuracy was 0.9229 (95% CI: 0.9099-0.9319) for syndrome pattern in test dataset with Corpus 1, and 0.9559 (95% CI: 0.9429-0.9699) for syndrome pattern in test dataset with corpus 2.

**Figure 4:**
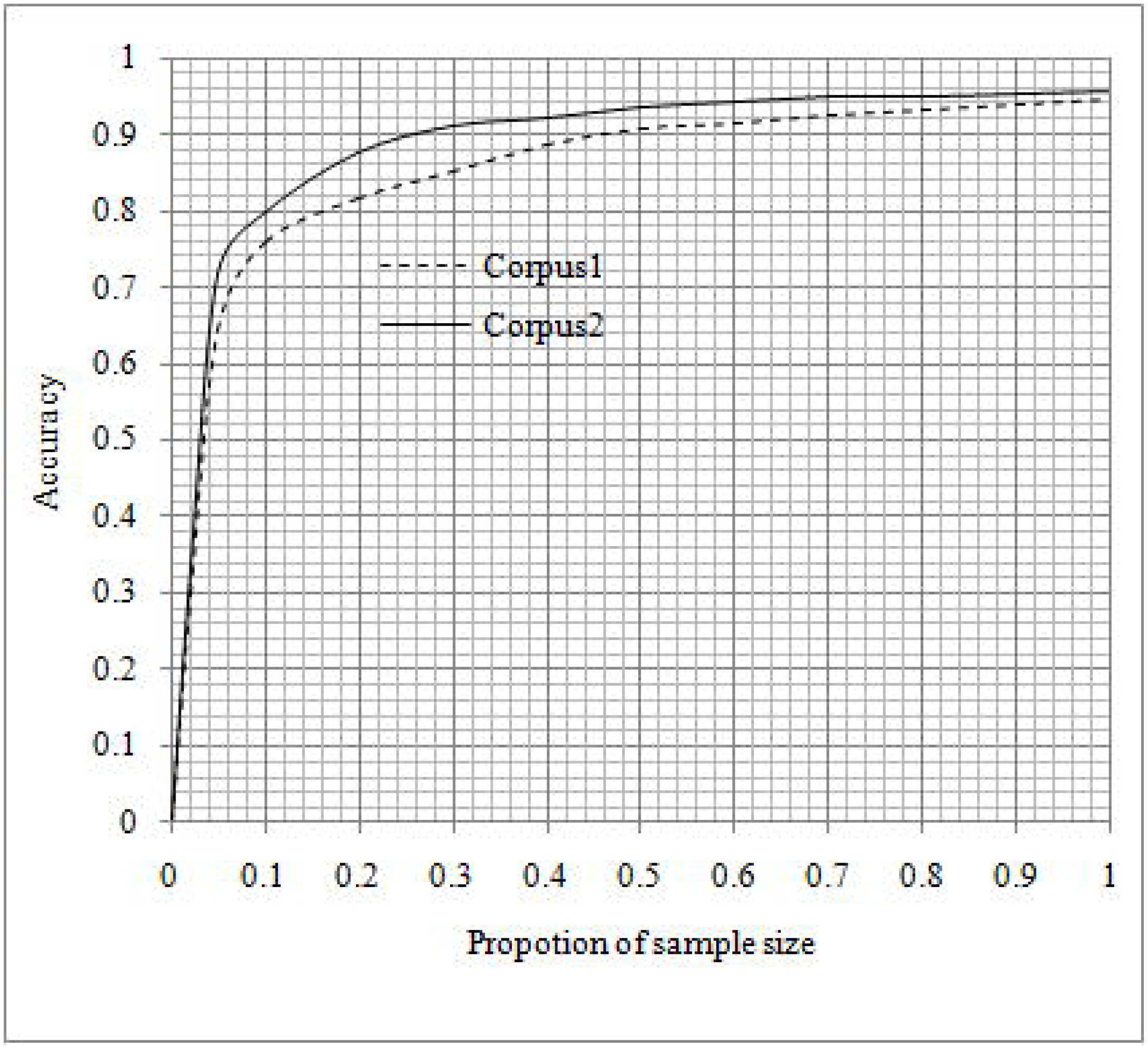
The comparison of performance on model-based reasoning methods on syndrome pattern diagnosis on integrative medicine of lung disease based on Corpus 1 and Corpus 2. Corpus 1 consisting of syndrome and sign information, Corpus 2 consisting of syndrome and sign information plus clinical diagnosis information.

### Association of accuracy and sample size with syndrome pattern type

We performed an average accuracy analysis in the development dataset classified by the number of syndrome pattern type and each group’s sample size. The results showed that the average accuracy increased with the increase in sample size of each group and decreased with the increase in number of syndrome pattern (Table 7). The linear regression analysis showed that each group’s sample size was significantly associated with the number of syndrome pattern with an accuracy of 0.90 (Y = 34.39×X+109.43, p<0.0001, Y: each group sample size, X: number of syndrome pattern type) and 0.95 (Y = 48.55× X + 296.78, p<0.0001, Y: each group sample size, X: number of syndrome pattern type), respectively (Figure 5).

**Table 7:** The average accuracy analysis grouped by sample size of each group and number of syndrome pattern type

| Each group sample size | N = 2 | N=3 | N=4 | N=5 | N=6 | N=7 | N=8 | N=9 | N=10 |
| --- | --- | --- | --- | --- | --- | --- | --- | --- | --- |
| 16 | 0.5714 | 0.4001 | 0.3876 | 0.3122 | 0.2521 | 0.3113 | 0.3076 | 0.2068 | 0.1875 |
| 40 | 0.6575 | 0.5001 | 0.4375 | 0.3511 | 0.2916 | 0.3751 | 0.3751 | 0.2916 | 0.2251 |
| 64 | 0.7238 | 0.6412 | 0.5384 | 0.5125 | 0.4636 | 0.4444 | 0.4174 | 0.4127 | 0.3921 |
| 80 | 0.8751 | 0.7291 | 0.6406 | 0.6311 | 0.5521 | 0.4732 | 0.5468 | 0.4513 | 0.4001 |
| 160 | **0.9375** | 0.8542 | 0.8437 | 0.8432 | 0.8345 | 0.7901 | 0.7621 | 0.7577 | 0.7325 |
| 240 | 0.9375 | **0.9097** | **0.9014** | **0.9011** | 0.8993 | 0.8482 | 0.8515 | 0.8487 | 0.8083 |
| 320 | **0.9658** | 0.9114 | 0.9074 | 0.9151 | **0.9227** | 0.8973 | 0.8984 | 0.8836 | 0.8515 |
| 400 | 0.9688 | 0.9433 | 0.9384 | 0.9281 | 0.9301 | **0.9266** | **0.9023** | **0.9025** | 0.8929 |
| 480 | 0.9752 | **0.9553** | 0.9414 | 0.9412 | 0.9418 | 0.9464 | 0.9444 | 0.9234 | **0.9135** |
| 560 | 0.9762 | 0.9583 | **0.9534** | **0.9521** | **0.9532** | 0.9482 | 0.9487 | 0.9394 | 0.9304 |
| 640 | 0.9776 | 0.9653 | 0.9633 | 0.9661 | 0.9626 | **0.9526** | **0.9619** | 0.9456 | 0.9354 |
| 720 | 0.9786 | 0.9708 | 0.9688 | 0.9712 | 0.9709 | 0.9672 | 0.9678 | **0.9591** | 0.9356 |
| 800 | 0.9813 | 0.9776 | 0.9756 | 0.9735 | 0.9739 | 0.9785 | 0.9734 | 0.9597 | 0.9429 |

Note: The first average accuracy arrived at 0.90 and 0.95 were bold.

**Figure 5:**
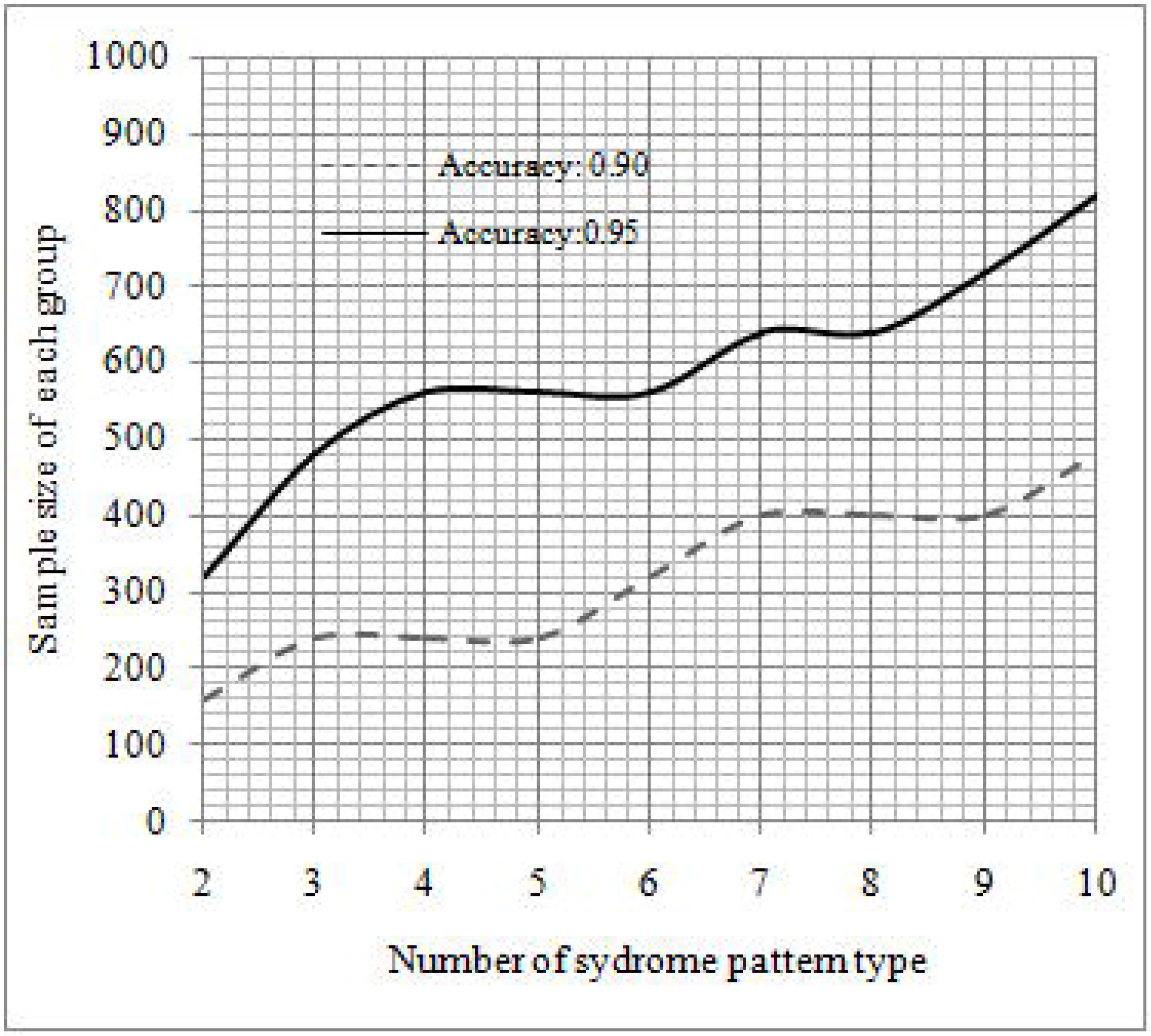
The association analysis between sample size of each group and number of syndrome pattern type at average accuracies of 0.90 and 0.95.

## Discussion

We developed MBR methods for diagnosis in integrative medicine in lung diseases based on a real-world EMR dataset with NLP. In this work, real-world medical records of clinical cases were used to develop models, and medical texts were mapped to vectors of real numbers that a computer could process. CNN approaches can automatically extract features from word vectors, thus contributing to the high performance of MBR methods in syndrome pattern diagnosis in integrative medicine in lung diseases. To the best of our knowledge, this study is the first to investigate MBR methods for diagnosis in integrative medicine on a large real-world dataset using NLP and deep learning methods in China. These MBR methods can be recommended for a clinical decision-making system and can also provide a novel approach for diagnosis in integrative medicine.

An interesting finding is the high performance of the MBR methods for syndrome pattern diagnosis in integrative medicine. The best Word2Vec CNN MBR in syndrome pattern diagnosis in integrative medicine had an accuracy of 0.9471 and 0.9250 in the development dataset and external dataset, respectively. Word embedding and CNN contributed to the high performance. Word embedding techniques can map texts to computability vectors, which can perform text analysis with quantitative analysis. CNN can automatically extract features from medical texts, significantly contributing to performance of the MBR. Additionally, the diagnosis information of modern medicine being added to the corpus enhances the accuracy of the syndrome pattern diagnosis in integrative medicine with reasoning, thus indicating that physicians can more efficiently make a syndrome pattern diagnosis after determining the diagnosis of modern medicine.

We performed an association analysis to evaluate the relationship between the number of syndrome pattern type and each group’s sample size for the accuracy of MBR algorithms. Moreover, we conducted a linear regression analysis to estimate the linear function of each group sample size and syndrome pattern type at an accuracy of 0.95. Only a few studies reported on the quantitative associations. In the Word2Vec CNN MBR algorithms at an accuracy of 0.95, the smallest group sample size was 300 for two syndrome pattern types, and each group sample size had at least 800 for 10 syndrome pattern types. According to the linear model, the Word2Vec CNN MBR based on each group’s sample size with at least 1,200 showed high performance in syndrome pattern with 20 types. A total of 400 common syndrome pattern types were grouped into 20 systems in integrative internal medicine. A total of 25,000 medical records of clinical cases could satisfy the word2Vec CNN MBR methods in syndrome pattern diagnosis in an integrative system at an accuracy of 0.95. A total of 500,000 medical records of clinical cases could satisfy the word2Vec CNN MBR methods in the diagnosis of 400 syndrome patterns in the entire integrative internal medicine at an accuracy of 0.95.

Interestingly, the MBR combined with RBR methods in syndrome pattern diagnosis in integrative medicine showed high performance. Specifically, Word2Vec CNN MBR combined with RBR methods had an accuracy of 0.9559 in syndrome pattern diagnosis in corpus 2 with additional information on modern medicine diagnosis. This reasoning method showed a more understandable and clearer knowledge of lung diseases for physicians in comparison with the Word2Vec CNN MBR methods. Moreover, it was more suitable for users of or physicians practicing integrative medicine. Generally, a hybrid reasoning is more suitable for application in clinical practice.

Although this study used novel methods to develop MBR in syndrome pattern diagnosis in integrative medicine, it has several limitations. First, we selected only10 out of the 20 common syndrome pattern types in lung diseases partly because the other 10syndrome pattern types did not have enough medical records of clinical cases. Therefore, future studies should use comprehensive syndrome patterns in lung diseases or other systems. Second, the size of the corpus for pre-trained word vectors was not large to cover all Chinese words or special items on lung diseases.

## Conclusion

MBR methods based on Word2Vec CNN showed high performance in syndrome pattern diagnosis in integrative medicine in lung diseases. The parameters of each group sample size, syndrome pattern type, and clinical diagnosis of lung diseases were associated with the performance of the methods.

## Data Availability

The datasets generated and/or analyzed during the current study are not publicly available due to private information but are available from the corresponding author on reasonable request. Dataset are from the study whose authors may be contacted at Center of Bioinformatics and Biostatistics, Institutes of Integrative Medicine, Fudan University. The data concerning external test dataset and an example of development of dataset were available in https://github.com/zihuitang/clincial_decision_support_system_im .

## Abbreviations

ANN: Artificial neural network
CI: Confidence interval
CNN: Convolutional neural network
EMRs: Electronic medical records
XGBoost: Extreme gradient boosting
KNN: K-nearest neighbor
MBR: Model-based reasoning
MLP: Multilayer perceptron
NLP: Natural language processing
RF: Random forest
RBR: Rule-based reasoning
SVM: Support vector machines
TCM: Traditional Chinese medicine

## Declarations

Ethics approval and consent to participate

The study was approved by Ethics Committee of the Huashan Hospital (approval number: HIRB-2018-166) and performed in accordance with the Declaration of Helsinki.

## Consent for publication

Not applicable.

## Patient and Public Involvement

With a real-world study design, electronic medical records on patient were used for diagnosis modeling based on artificial intelligence. All data and attributes were extracted from electronic medical records with data analysis approaches.

## Author’s Contributions

W.G and X.Q drafted the manuscript. Z.W and Q.K participated in the design of the study and performed the statistical analysis. Z.T and L.J conceived of the study, and participated in its design and coordination and helped to draft the manuscript. All authors read and approved the final manuscript.

## Acknowledgments

We thank the grant from Institutes of Integrative Medicine of Fudan University to support the study.

## Authors’ Information

W.G, Q.K, Z.T, and L.J were Department of Integrative Medicine, Huashan Hospital, Fudan University, Shanghai, China; X.Q was Department of Neurosurgery, Fudan University, Shanghai, China; Z.W was Shanghai Sunjian Informatics Technology Company Limited, Shanghai, China.

